# Health workers Motivators to uptake of the Covid-19 vaccine at Iganga Hospital Eastern Uganda, and Mengo Hospital Kampala Uganda; A qualitative study

**DOI:** 10.1101/2021.10.25.21265494

**Authors:** Lubega Muhamadi, Namulema Edith, Waako James, Nazarius Mbona Tumwesigye, Safinah Kisu Museene, Stefan Swartling Peterson, Anna Mia Ekström

## Abstract

Uganda continues extensive mobilization and administration of the Covid 19 vaccine to its people albeit some vaccine hesitancy with in the population. Amongst the health workers however, approximately 70% had received their first dose while 40% had received their second dose of the Covid-19 vaccine by September 2021 respectively. These figures represent a recognizable acceptance rate among health workers. Exploring motivators to vaccine uptake among health workers is vital for the government’s general population vaccine rollout plan.

We conducted 12 focus group discussions and 20 in-depth interviews with health workers (vaccinated and unvaccinated) to understand motivators to vaccine acceptance in their own perspective in central and eastern Uganda. Reported motivators to vaccine acceptance included; risk susceptibility/protection, fear of death and/or cost of treatment and experiences of Covid related grief. Other were trust in the vaccine, call to government policy and vaccine success stories elsewhere, real or perceived benefits of vaccination and peer influence.

We recommend intensified dissemination of health worker tailor made tools/guides for information, education and communication about the Covid 19 vaccine. The tools need to emphasize the elicited themes/motivators. We also recommend use of peers who have taken up the vaccine and survived Covid-19 or got a mild form of the disease to elicit positive peer influence about the vaccine amongst health workers. The information dissemination and peer narratives could be done through the health worker’s social media platforms, union or association websites, personal statements, editorials or other media.

## 1.0 Background

The government of Uganda like many other parts of the world continues extensive mobilization and administration of the Covid 19 vaccine to its people albeit the prevailing vaccine hesitancy with in the population (MOH 2021, Museveni 2021). The government projects vaccination of approximately 12 million people before December 2021. The first phase of the vaccination rollout started in March 2021 and targeted to vaccinate a population of 4.8 million people. The targeted categories included; health workers, security personnel, people above 50 years, below 50 years but with underlying co-morbidities, teachers and students aged 18 years and above by June 2021 (MOH 2021, MOH 2021, Museveni 2021). Unfortunately, the country still faces a hindrance of vaccine hesitancy among this target population. By September 2021, 81% of the targeted 4.8 million people had not received their first dose, while 91% had not received their second dose of the vaccine (Museveni 2021). The vaccine acceptance rate however varied amongst the different categories of the target population with 5.5% (people with comorbidities) being the lowest and 74.9% (health workers) being the highest, measured by 1^st^ dose coverage (Museveni 2021). Amongst the health workers who are the most vulnerable frontline staff in the fight against Covid-19, approximately 70% of the health workers had received their first dose while 40% had received their second dose of the Covid-19 vaccine by September 2021. These figures represent a recognizable acceptance rate among health workers albeit some hesitancy compared to other categories among the target population (MOH 2021).

Health workers are trusted people and communities follow their example and health actions almost religiously. Therefore as the government vaccine roll out plan evolves, understanding what motivates them to accept the vaccine is likely to positively impact how the general community takes up the vaccine as well (Bashirian, Jenabi et al. 2020, Wang, Ahorsu et al. 2021). A few online surveys indicated that self-protection; belief in science, stopping the virus, age, education among others motivated communities to accept the vaccine (Agha, Chine et al. 2021, Dodd, Pickles et al. 2021, Wang, Ahorsu et al. 2021). There is, however, paucity of information on health workers motivators for uptake of the Covid vaccine since the vaccination started in Uganda. An in-depth exploration of the health workers perceptions towards this vaccine from their own perspective and context is therefore vital especially at a time when the government is rolling out vaccination to over 20 million people and hence this study.

## 2.0 Materials and Methods

### Study Setting and population

Between June and August 2021, we sought to explore health workers perceptions with regard to the Covid-19 vaccine in Mengo and Iganga hospitals. Mengo hospital is a private not for profit entity located in the capital city Kampala. The hospital offers both general clinical and specialized services to an urban population of over three million people from the Kampala metropolitan area. The hospital employs over 800 staff consisting of technical, administrative and support staff. It is one of the designated Covid-19 treatment centres in Kampala offering both general clinical care and supportive treatment to people infected with Covid-19. It also has a high dependency unit (HDU) for clients that need high level monitoring and an intensive care unit (ICU) for critical cases. Over the first and second wave of the Covid-19 pandemic in Uganda, the hospital treated over 300 patients of different levels of severity of the infection. The hospital also serves as a Covid-19 vaccination centre (Director 2021). Iganga hospital is located in the east of Uganda approximately 120 km east of the capital city Kampala. It is a public general referral hospital offering both general clinical and specialized services. The hospital is a referral centre for about six districts constituting a population of about three million people. Most of the patients here are predominantly rural living on subsistence farming with only approximately 7% living in urban and peri-urban environments. The hospital has approximately 200 staff consisting of technical, administrative and support staff. It is one of the designated Covid-19 treatment centres in the region offering both general clinical care and supportive treatment to people infected with Covid-19 but also has a HDU. Over the first and second wave of the Covid-19 pandemic in Uganda, the hospital treated over 200 patients of different levels of severity of the infection, fifteen of whom were health workers. The hospital also serves as a Covid-19 vaccination centre (UBOS 2014, DHO 2021, MOH 2021).

This qualitative study employed 12 focus group discussions (FGDs) comprising staff of different cadres and deployment. Six of the FGDs were conducted in Mengo hospital while another six were conducted in Iganga hospital. We also conducted 20 in depth interviews (IDIs) with health workers from both Mengo and Iganga hospitals 10 of whom had been vaccinated with the Covid-19 vaccine and 10 had not been vaccinated. The study aimed at exploring possible motivators/barriers for uptake of the vaccine amongst the health workers from their own perspective (Boyden 1997, Holloway 2008). The FGD and IDI respondents were purposively selected based on maximum variety sampling from a sampling frame covering all levels of cadres from specialist health personnel to the lowest support staff in order to elicit different viewpoints. The respondents were selected because they were considered to be more “knowledge rich” on the study subject in their own situations than anybody else (Boyden 1997, Fleury and Lee 2006, Stewart DW, Shamdasani PN et al. 2007, Holloway 2008). The participants were chosen from consenting members in both hospitals. The FGDs were stratified by cadre to encourage more active participation and included those who had been vaccinated and those who had not been vaccinated to avoid potential stigmatization. Participants were invited to take part in only one FGD or IDI. Each FGD consisted of a maximum of 12 participants. While each of the participants knew their vaccination status, the participants were not told how their groups were selected or their vaccination status to avoid “playing to type”

### Data collection tools and methods

Using a topic guide, the selected participants were probed on their knowledge, attitudes and perceptions about the Covid-19 vaccine. Their views on values and norms related to risk/benefit of vaccines and public health interventions, acceptance and uptake of vaccines in general and the Covid-19 vaccine in particular, as well as prevailing common misconceptions and myths about the vaccine among others were also probed. Interviews were stopped when it was judged that saturation had been reached and no more new information could be retrieved from the respondents. All data collection was supervised and assessed by the first author LM) who is a male, indigenous public health physician and the second and second third authors (NE WJ,) both of whom are conversant with qualitative research and the health system dynamics for both Iganga and Mengo hospitals respectively. Five research assistants moderated and took notes for the study. The research assistants were conversant with qualitative data collection methods, had conducted similar research in the study settings and were all fluent in Luganda and Lusoga (the local languages). They were trained for two days on the study aim, design and tools. Role-plays were used to prepare the research assistants for their interaction with the informants (Dahlgren and M. Emmelin 2004, Fleury and Lee 2006, Henderson and Naomi R 2009). The experiences from the role-plays were discussed and further methodological guidance was given.

### Data management and analysis

All the FGDs and the IDIs were conducted in a mixture of English, Lusoga or Luganda (respective native languages), transcribed and later all translated into English by the interviewers. The authors listened to the audio recordings to confirm the validity of the information. Data collection stopped when information relating to the topic guides revealed no new information. Data analysis was iterative including reviews and discussions at different stages of data collection and appropriate modifications were made in the tools to address emerging issues (Morse F 1995, Stewart DW, Shamdasani PN et al. 2007). The units of analysis were the transcripts from the IDIs and the FGDs. Content analysis was used to analyze the transcripts. This entailed reading and reviewing texts of the entire interview back and forth to identify meaningful units in relation to the study subject (Morse F 1995, Dahlgren and M. Emmelin 2004, Holloway 2008). The meaningful units were condensed and coded by categories and themes, and discussed by (LM, NE and WJ and) until consensus was reached on the appropriate codes and the themes (Dahlgren and M. Emmelin 2004).

### Ethical clearance

The study was approved by the Mengo hospital Research and Ethics Committee (REC) ref MH/REC/39/06-2021, and the Uganda National Council for Science and Technology (UNCST). We also sought the approval of the management of both Mengo and Iganga hospitals. As part of the informed consent, the participants were told about the aims of the study, the anticipated benefits and risks, their ability to participate or withdraw at any time, and assured that all information obtained would be kept confidential. The participants signed two copies of the consent form before the interview commenced, and one copy was given to the participant.

## 3.0 Results

**Table 1:**
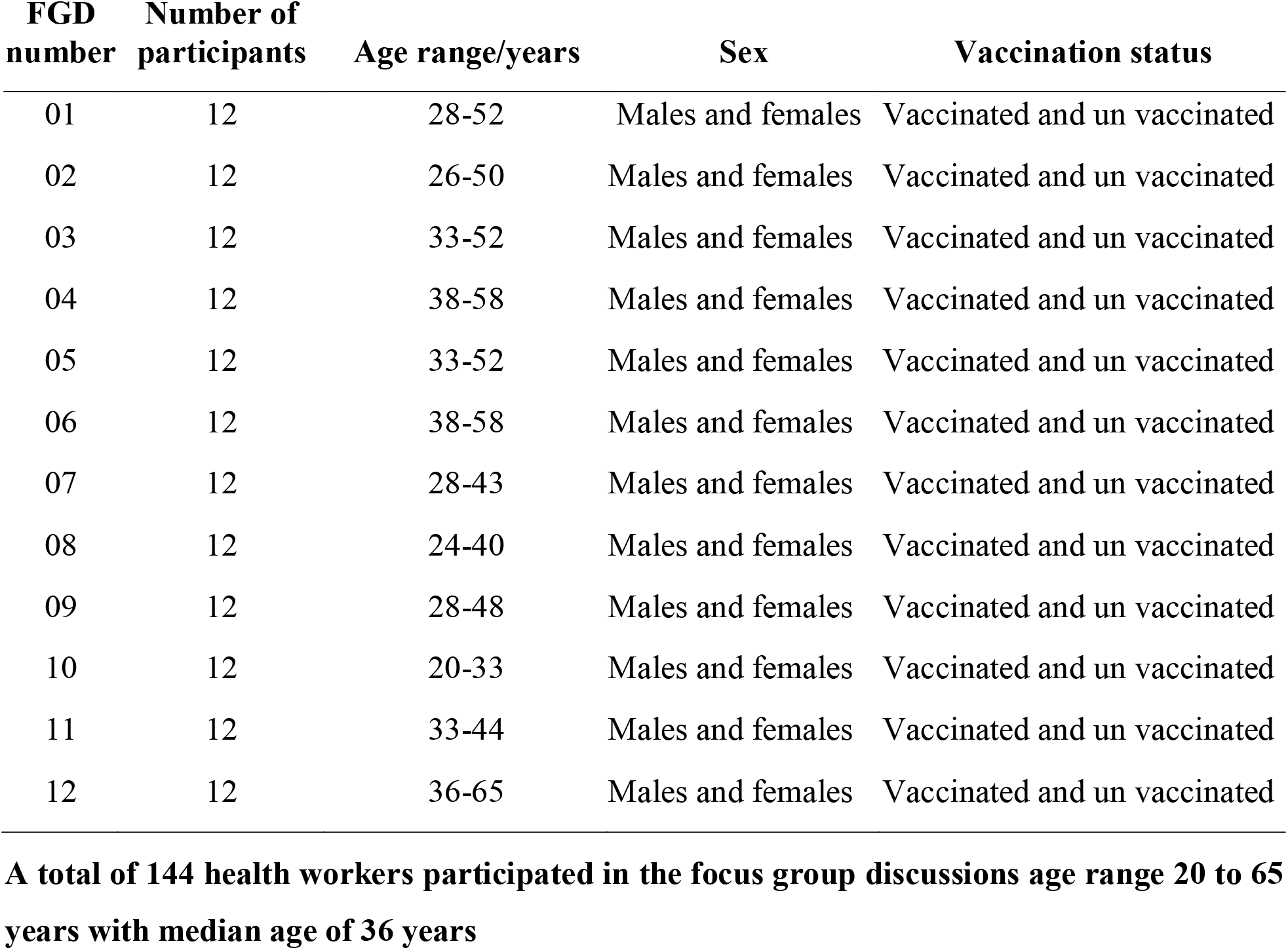
Summary characteristics of the FGD participants stratified by cadre or unit of health care.

**Table 2:** Summary Characteristics of the IDI participants purposively identified by cadre.

| **Participants number** | **Age range/years** | **Sex** | **Vaccination status** |
| --- | --- | --- | --- |
| 01 | 31-35 | Male | Vaccinated |
| 02 | 31-35 | Female | Vaccinated |
| 03 | 46-50 | Male | Un vaccinated |
| 04 | 36-40 | Male | Un vaccinated |
| 05 | 36-40 | Male | Un vaccinated |
| 06 | 41-45 | Female | Vaccinated |
| 07 | 36-40 | Female | Vaccinated |
| 08 | 31-35 | Male | Unvaccinated |
| 09 | 26-30 | Male | Un vaccinated |
| 10 | 26-30 | Male | Vaccinated |
| 11 | 36-40 | Female | Vaccinated |
| 12 | 31-35 | Female | Un vaccinated |
| 13 | 36-40 | Female | Un vaccinated |
| 14 | 26-30 | Female | Vaccinated |
| 15 | 26-30 | Female | Vaccinated |
| 16 | 31-35 | Female | Vaccinated |
| 17 | 46-50 | Male | Un vaccinated |
| 18 | 46-50 | Female | Un vaccinated |
| 19 | 31-35 | Female | Un vaccinated |
| 20 | 51-55 | Female | Vaccinated |

**Twenty participants were identified for the in depth interviews. Twelve were females and 8 males. Age range was 28 to 55 years with a median of 36 years.**

Our analysis of textual data generated seven themes including: (1) risk susceptibility and protection (2) fear of death and/or cost of treatment, (3) experiences of Covid related grief (4) trust in the vaccine (5) call to government policy and vaccine success stories elsewhere, (6) real or perceived benefits of vaccination and (7) Peer influence.

### Risk susceptibility and protection

Health workers reported that they took the Covid vaccine because they saw themselves at a greater risk of contracting Covid given they were working on Covid wards or in other hospital wards that exposed them to Covid patients. Others reportedly took the vaccine as a precaution for personal protection and protection of their families and immediate social networks.

> “*What made me get vaccinated was that us here who work in the hospital, we get exposed to many Covid patients and some patients whose Covid status we don’t even know. In that situation and the other kinds of work we usually do, we are never safe. That is what prompted me to go and get vaccinated and even the nature of the wave, the way the rate of the disease was, I was like if there is any chance for me to get vaccinated, let me go and I get vaccinated. “Asiika obulamu ttasa muka” meaning that you don’t joke around when you are saving life, so I went and I had my first dose and the second one” (male informant, vaccinated)*.

> “*This disease is so infectious, so some of us with families thought it a good idea to get vaccinated so we can protect our families and those other people around us. It was a fantastic chance to miss” (female informant, vaccinate)*.

### Fear of death or cost of treating Covid

Fear of death caused by or related to Covid given its high attack, transmission, and fatality rates was another motivator for some health workers accepting the vaccine. Closely associated was the fear of the high cost of treatment (direct and indirect) for the disease which convinced the health workers to vaccinate as a cheaper preventative measure compared to care and treatment of Covid.

> “*This Covid disease is so easily transmitted and has killed so many people in a short period of time……in addition the cost of treatment is so high. Even with free treatment in the hospital, you get hospitalized for so long, lose work and income among many other things so some of us found it cheaper to get the vaccine than wait for the disease, get bed ridden or even death, we have seen this with our own eyes” (female informant, vaccinated)*.

### Experiences of Covid related grief

Some health workers had seen their friends at work and other Covid patients die of Covid with in their own hands of care. Others had lost family members, professional colleagues, or friends with in their social networks in unbearable grief. These workers therefore saw the chance to get the Covid vaccine when available as golden and took it with a sigh of relief.

> “*Well I could say that in my family, we lost four people to Covid, myself I can tell you that I lost my best man together with his wife who was also the matron. I have a colleague, a friend of mine that I stayed with during my graduate studies, he died……*.*even here at the hospital, many of our workmates have succumbed to Covid. So I think you can see the effects it has had on me and same story to many of us. So when the chance to get the vaccine came I grabbed it quicly with both hands” (male informant, vaccinated)*.

### Trust in the vaccine

Other respondents reported trust in the vaccine as a motivation for them accepting to take it up. Such respondents trusted the production processes of the vaccine, its efficacy given that it had been qualified by the WHO and the Ministry of Health together with previous UNEPI and government’s successful and safe previous vaccination programs in the country.

> “*Personally I am confident about the vaccine. It works otherwise the WHO and the government would not have accepted it. Which government would administer a dangerous vaccine to its people……*.*these vaccines go through proper scrutiny before they are allowed for use. Besides, other UNEPI and government immunisation programs in Uganda have been very effective, so why doubt this one” (female informant, vaccinated)*.

### Call to government policy and vaccine success stories elsewhere

Health workers were reportedly motivated to take up the vaccine because of the sustained governments call for them to do so as a preventive measure against escalation of the pandemic. This call together with vaccine success stories in other countries like the United states, and Britain that had been hardly affected by Covid but were succeeding in stopping the pandemic through vaccination gave them optimism to accept the vaccine

> “*The government tells us almost every day that they have given us the vaccine because they don’t want us to get infected with covid-19 or severe Covid and get near intensive care unit and that it is also the best way to stop the pandemic. Even countries like the United States and Britain have managed to control the pandemic only through vaccination, these countries were badly hit by Covid but have now gotten back to normal because of vaccination so why not Uganda too” (female informant, vaccinated)*

> “*What prompted me, I went for training as a vaccinator at Imperial hotel and I was well educated about the importance of taking the vaccine so I took one myself………by the way after this education, even the negative perceptions and myths about the vaccine that I had heard before cleared because I got to know that the vaccine works and it is safe” (male informant, vaccinated)*

### Real or perceived benefits

Real or perceived benefits of getting Covid vaccination were often reported as motivators for health workers taking up the vaccine. The benefits reported included; being able to get a vaccination certificate which would be paramount for future travels outside the country, job security, returning to work, attending social functions and a vehicle to social fitting and respect. Other health workers reportedly saw taking up the vaccine as the only way that government would eventually lift the country lock down and opening of schools for their children.

> “*In my view, one has to have a vaccination certificate if they are going to retain their jobs, go back to work after the lock down, travel outside the country or attend any social fitting. It is going to be a requirement like a national identity card………….but also the truth is according to the president the only way government is going to lift the lockdown or open schools for our children is when many people get vaccinate. Surely that should be enough for anybody to seek vaccination as soon as the chance comes” (male informant, vaccinated*).

### Peer influence

Some health workers reported they had been motivated to take up the vaccine by peers at the same work place or elsewhere who had taken the vaccine with minimal or no side effects.

The reassurance from these peers swept any cast of doubt about the safety, efficacy and effectiveness of the vaccine and hence the energy for them to get vaccinated.

> “*We have friends here or in other places of work who took up the vaccine while we lay in fear but eventually most of them did not get any serious side effect apart from slight headache. So they talked to us and convinced us to do the same because it was safe and effective to them………actually we later realized that most of us who had gotten the vaccine became safe from Covid or in worst cases got mild forms of it” (male informant, vaccinated)*

## 4.0 Discussion

Our findings indicate that motivators for health workers accepting the Covid 19 vaccine were a function of multiple cross cutting themes including; risk susceptibility and protection, fear of death and/or cost of treatment and experiences of Covid related grief. Other themes were trust in the vaccine, call to government policy and vaccine success stories elsewhere, real or perceived benefits of vaccination and peer influence.

Risk susceptibility and personal protection/protection of family are functions of awareness, belief and appropriate knowledge the health workers had about how exposed they were to the Corona virus by virtue of their occupation. The health workers saw, treated Covid patients or even witnessed others under their care die of the illness. They appreciated how easily transmittable, expensive to treat in terms of direct and indirect costs and fatal the virus was and recognized they were at risk of contracting Covid. Their willingness to get vaccinated as a measure for prevention of getting Covid and or transmitting it to their families/social networks amplifies their knowledge scope about the importance of taking up the vaccine. Relatedly, some health workers had lost family members or friends due to Covid, the loss and grief caused by these deaths could have had a pushing effect for them to get vaccinated and avoid death. Risk susceptibility/ risk perception as well as personal protection and protection of others as drivers for vaccine acceptance has been established in other vaccine related studies in high and low income countries (Godinot, Sicsic et al. 2021, Guidry, Laestadius et al. 2021, Huynh, Tran et al. 2021, Viswanath, Bekalu et al. 2021, Wong, Wong et al. 2021). Fear of death, serious effects of the disease or experience of losing a loved one as a motivator for health intervention uptake has been established in other studies as well (Kim, Han et al. 2020, Abdulmoneim, Aboelsaad et al. 2021, Karlsson, Soveri et al. 2021).

Some health workers heeded to the sustained government appeal for them to get vaccinated to prevent severe Covid, had trust in the vaccine and attributed several benefits to getting vaccinated against Covid 19. This social phenomenon could be explained by the government’s sustained efforts in explaining the benefits against risks of getting the vaccine and thrusting community trust and confidence in the vaccine. Dissemination of appropriate information, education and communication (IEC) about the vaccine by the government positively impacted health workers to take up the vaccine. The government’s efforts in IEC dissemination likely generated belief, positive perceptions and attitudes towards the vaccine. The government’s insistence on vaccination certificates/mandate for all the vaccinated as well generated positive benefits (perceived or real) which motivated a few health workers to accept the vaccine. The success stories of the vaccine roll out in other countries as well had a positive impact on belief, attitude and perceptions of the health workers towards taking up the vaccine. The importance of dissemination of appropriate IEC materials on benefits/risks of health interventions as drivers for accepting health interventions and trust in a vaccine as a motivator for uptake has been established in other studies (Wilpstra 2020, Abdulmoneim, Aboelsaad et al. 2021, Binub and Haveri 2021, Dodd, Pickles et al. 2021, Kumari, Ranjan et al. 2021, Ojikutu, Stephenson et al. 2021, Osur, Chengo et al. 2021, Urairak 2021). Health intervention success stories as drivers for uptake of an intervention is also amplified in literature (Ramey, Downing et al. 2008, Seifert, Chapman et al. 2012, Chew, Sim et al. 2019, Wood and Schulman 2021)

Some health workers were motivated by the positive experiences from their colleagues who had been vaccinated but had minimal or no side effects and yet still did not suffer from Covid or severe Covid at all. Human behavior is not a function of only their genetics, knowledge, attitudes or perceptions but also influenced by among others their immediate family, friends or social networks (Fleury and Lee 2006). Individuals are most likely to accept/ refuse health events about their health depending on how much their families or friends think or have gone through regarding the same phenomenon as has been amplified by other health models or studies (Fleury and Lee 2006, Kumar, Quinn et al. 2012, Gruenewald, Remer et al. 2014, Golden, McLeroy et al. 2015, Kolff, Scott et al. 2018)

## 5.0 Conclusions/Recommendations

The government should consider intensified dissemination of health worker tailor made tools/guides for information, education and communication about the Covid 19 vaccine. The tools need to emphasize the importance of risk susceptibility to Covid and vaccination for self and community Covid prevention for all health workers. The tool should as well highlight the benefits versus risks for accepting the Covid vaccine, instigate more trust for the vaccine, the high treatment costs for Covid when one gets infected as well as the avoidable case fatalities of Covid.

Peers who have taken up the vaccine and survived Covid-19 or got a mild form of the disease could also be used to elicit positive peer influence about the vaccine amongst health workers. The information dissemination and peer narratives could be done through the health worker’s social media platforms, union or association websites, personal statements, editorials or other media. Increasing awareness, information, education and communication about health interventions has been found to improve their uptake in several intervention models (Mo, Wong et al. 2019, Cocchio, Bertoncello et al. 2020). Positive peer influence has also been found useful in improving service uptake in many other interventions (Stilgoe and Cohen 2021).

## Study limitations

As it is with qualitative studies, our deductions are purely based on narratives from the respondents with no statistical inferences. The fact that the interviewees themselves lived this life and volunteered the information, however, strengthens the concept that the data derived from the interviews was objective to the topic of the study. Additionally, we also triangulated our data collection methods (FGDs, IDIs) and conducted our analysis iteratively. This helped us to check for consistency and contradictions inside and across the groups and interviewees. Also, our team was multi-disciplinary, grounded in the collection and analysis and had a good contextual understanding of aspects relating to uptake of the Covid-19 vaccine in the study settings. We therefore feel that the content analysis employed for this study has achieved appropriate in-depth analysis for the purpose of the study.

## Data Availability

All data produced in the present study are available upon reasonable request to the authors

